# Weight Gain Following a Diagnosis of Anti-neutrophil Cytoplasm Antibody-Associated Vasculitis

**DOI:** 10.1101/2024.05.29.24308107

**Authors:** Tania Salehi, Thomas French, Tariq E Farrah, Neeraj Dhaun, Robert W Hunter

**Author notes:** **Corresponding Author:** Tania Salehi Clinical Fellow in Renal Medicine Royal Infirmary of Edinburgh.

## Abstract

**Objectives:** Patients with anti-neutrophil cytoplasm antibody (ANCA)-associated vasculitis (AAV) are at increased long-term risk of cardiometabolic diseases. The prevalence of obesity in AAV has not been well documented. We aimed to characterise change in body weight following a diagnosis of active AAV and to determine the risk factors for this.

**Methods:** We examined data from a single-centre registry of patients with AAV, diagnosed between 2003 and 2023. We evaluated changes in body weight and BMI following diagnosis. Using linear regression, we identified factors contributing to an increase in BMI at six-months. Logistic regression was used to define predictors for obesity at six-months.

**Results:** Two-hundred and fifteen patients with active AAV were included in the analysis. Patients experienced a mean gain in body weight of 5.2% in the first six-months; this was maintained for at least two-years. 64.1% of patients were overweight or obese at six-months. Weight gain was greater following first presentation of AAV compared to relapsing disease. Baseline factors associated with an increase in BMI at six-months included higher eGFR (β=0.70 [0.36-1.03], P<0.001) and earlier year of presentation (β=0.38 [0.08-0.69], P=0.008). Higher eGFR (aOR=1.36 (1.08-2.72), P<0.001) and baseline BMI (aOR=2.57 (1.81-3.64), P<0.001) were associated with an increased likelihood of obesity at six-months.

**Conclusion:** Weight gain is common following a diagnosis of active AAV. This is less pronounced than it was two-decades ago. Better kidney function and higher baseline BMI are associated with a greater risk of being obese at six-months. Management of AAV should include risk mitigation for developing an unhealthy high BMI.

**Key messages:**

1. Weight gain and an unhealthy high BMI are prevalent following diagnosis of active AAV.
2. Higher baseline eGFR is associated with greater weight-gain in the first six-months following diagnosis.
3. Weight gain is less pronounced following treatment of relapsing disease compared to initial presentation.

## Introduction

Anti-neutrophil cytoplasm antibody (ANCA)-associated vasculitis (AAV) is a rare, potentially life-threatening autoimmune disease characterised by necrotising inflammation in small vessels, causing end-organ injury [1, 2]. The management of severe AAV involves the use of combination immunosuppression including high-dose glucocorticoids, cyclophosphamide and/or B-cell depletion therapy. Short-term mortality has fallen substantially as treatments have improved in recent decades. Thus, the major challenges facing individuals with AAV relate to the longer-term sequalae of organ damage and immunosuppressive treatments, including increased rates of cardiometabolic disease [3].

An increase in body weight following a diagnosis of active AAV has been observed in clinical trial populations. *Post-hoc* analyses of the WGET (Wegener’s Granulomatosis Etanercept) and RAVE (Rituximab in AAV) trials demonstrated that AAV patients treated with high-dose glucocorticoid regimens experienced significant gains in body weight in the first 6-12 months after diagnosis [4, 5]. However, in ‘real-world’ epidemiological studies, patients with AAV do not exhibit increased rates of obesity (whereas they are at increased risk of cardiovascular disease, diabetes mellitus, thyroid disease, osteoporosis, and venous thromboembolism) [6-8]. Thus, there is uncertainty around the extent of this potential problem outside clinical trial settings.

Understanding the prevalence of, and risk factors for, obesity in AAV would allow personalisation of treatment regimens to minimise risk of AAV-associated obesity and its complications. Thus, we characterised changes in body weight following a diagnosis of active AAV in a contemporary, real-world cohort, and determined the risk factors for this. In a secondary analysis, we determined whether obesity modifies risk of disease relapse, given that obesity and the metabolic syndrome are known to induce a pro-inflammatory state [9, 10].

## Methods

### Study design and population

We analysed data from a single centre registry of all patients presenting with active AAV between 2003 and 2023. “Active” disease was classified based on a combination of clinical, laboratory, radiologic and histological findings suggesting vasculitis disease activity, with exclusion of alternative causes, which required immunosuppressive treatment. In addition to PR3- and MPO-positive patients, those with ANCA-negative pauci-immune vasculitis were included in the analysis. We analysed routinely collected clinical and laboratory data retrieved from the electronic health record using SQL-based database queries. Our study analysed all data from vasculitis clinic visits as well as any other blood tests taken in primary or secondary care in our health board.

### Patient characteristics

At time of diagnosis of active AAV we recorded: age, sex, weight, body mass index (BMI), diabetic status, lipid profile, serum creatinine, estimated glomerular filtration rate (eGFR), urine protein:creatinine ratio (uPCR), and ANCA serology. During the treatment and follow-up phases we recorded: induction therapeutic agent(s), glucocorticoid exposure and relapse history.

The baseline weight was taken as the measurement closest to the date of AAV diagnosis, accepting any value within a window extending 90 days preceding and 30 days following diagnosis. Weight was recorded at every follow up visit to the vasculitis clinic. Height measurements were presumed to have remained unchanged throughout the study. In the event of multiple, discrepant height recordings for one individual, the median value was taken. BMI was classified into four subcategories corresponding to WHO definitions [11]; underweight: BMI less than 18.5 kg/m^2^, healthy weight: BMI between 18.5 and 24.9 kg/m^2^, overweight: BMI between 25.0 and 29.9 kg/m^2^; obese: BMI ³30.0 kg/m^2^.

Baseline diabetes and hyperlipidaemia were inferred from blood test data. A diagnosis of diabetes was assumed if there was a recorded haemoglobin A1c (HbA1c) level of ≥48 mmol/mol and/or random blood glucose level of ≥11.1 mmol/L at or prior to diagnosis. Patients were deemed to have hypercholesterolaemia in the presence of an elevated low-density lipoprotein cholesterol (LDL-C) of >3 mmol/L and/or total cholesterol >5 mmol/L at or prior to diagnosis. eGFR was calculated using the CKD-EPI equation [12]. ANCA status was classified as PR3 positive, MPO positive, dual PR3 and MPO positive or ANCA negative, according to the highest recorded value for PR3 and MPO ELISA titre at any time during follow-up. Induction immunosuppression was classified as being cyclophosphamide monotherapy, rituximab monotherapy, combination cyclophosphamide and rituximab or mycophenolate mofetil monotherapy. The doses of glucocorticoids used were documented both at the beginning of treatment (week 0) and at week 12, and were reported as prednisolone equivalents.

Remission was defined as clinically silent disease in a patient taking no more than 7.5 mg prednisolone per day. Relapse was defined as a change in symptoms or signs that—after appropriate investigation—was likely to be attributable to vasculitis disease activity and not to alternative pathology such as infection. In almost all instances this prompted an escalation in immunosuppression. All diagnoses of “active” AAV, “relapse” and “remission” were made by a Consultant Physician with experience of managing AAV patients.

### Endpoints

Our primary analysis determined how body weight and BMI change following a diagnosis of active AAV. We evaluated the trajectory of body weight over time post-diagnosis and initiation of immunosuppression for the index AAV presentation and after the first episode of disease relapse. We sought to identify baseline risk factors for a rising BMI and obesity at six months. In an exploratory secondary analysis, we examined whether obesity was associated with differences in rates of AAV relapse.

### Statistical analysis

Continuous variables were reported as group means ± standard deviation (SD) if normally distributed or as median ± interquartile range (IQR) if not.

The average relative differences in baseline characteristics between PR3 and MPO positive cohorts were compared by two-tailed t-tests for normally distributed continuous variables, Mann-Whitney-U tests for non-normally distributed continuous variables and chi-square testing for categorical variables. Paired two-tailed t-tests were used to evaluate changes in weight over time.

Ordinary least squares regression was used to identify predictors of percentage change in BMI at six months. In order to account for the non-linear relationship between age and change in BMI, a quadratic term was included in regression formulae concerning this variable (age + age^2^). Unadjusted regression coefficients were calculated through univariate linear regression, and adjusted regression coefficients were calculated adjusting for age, BMI at baseline, eGFR, induction agent, glucocorticoid dose at week 0, and year of presentation. These confounders were chosen using backwards stepwise regression, using the Akaike Information Criterion (AIC) as a means of model comparison. In order to approximate a normal distribution in the dependent variable (BMI percentage change at six months), seven outliers (identified by a distance from the median of more than 1.5 times the IQR) were not included in the final analysis. Assumptions of normality and heteroscedasticity were assessed using a Q-Q plot of residuals, and a predicted/observed scatter plot of residuals.

Univariate and multivariate logistic regression were used to calculate odds ratios (OR) for obesity at six months from induction therapy. Adjusted OR (aOR) were calculated adjusting for age, sex, baseline BMI, eGFR, ANCA sub-group, induction agent, glucocorticoid dose at week 0 and at 12 weeks, and year of presentation.

Time to event analysis was performed to identify risk factors for disease relapse. Time to either relapse or censorship from death or loss to follow up was used to generate survival tables. Cox proportional hazards regression was used to generate hazard ratios adjusted for age, sex, gender, ANCA subgroup and induction agent. Kaplan-Meier curves were compared with a log-rank test.

Missing data were treated as missing completely at random (MCAR), and regression analyses were completed on a ‘complete case’ basis. R Statistical Software (v4.3.2; R Core team 2023) was used for statistical analysis, along with the packages ‘readxl’, ‘plyr’, ‘tidyr’, ‘pROC’, ‘survival’, ‘ggsurvfit’ and ‘car’.

## Results

### Baseline characteristics

During the study period a total of 436 patients were diagnosed with first presentation AAV, of whom 215 had available baseline BMI data and were included in the analysis. The vast majority of those with missing BMI data had no recorded height. Patients were followed up for a median of 5.7 +/- (2.4-9.4) years.

Baseline demographic, clinical and laboratory characteristics of the cohort are presented in **Table 1**. Baseline BMI was 26.0 (22.9-30.1) kg/m^2^. Sixty-eight patients (31.6%) were overweight and 52 patients (24.2%) were obese at presentation with AAV. In nine patients, the induction immunosuppression regimen did not fall into our pre-specified categories (and was therefore not classified). Fifty-two patients (24.2%) experienced one or more clinical relapses during the follow-up period. Six patients (2.8%) had refractory disease and did not achieve clinical remission.

**Table 1.**
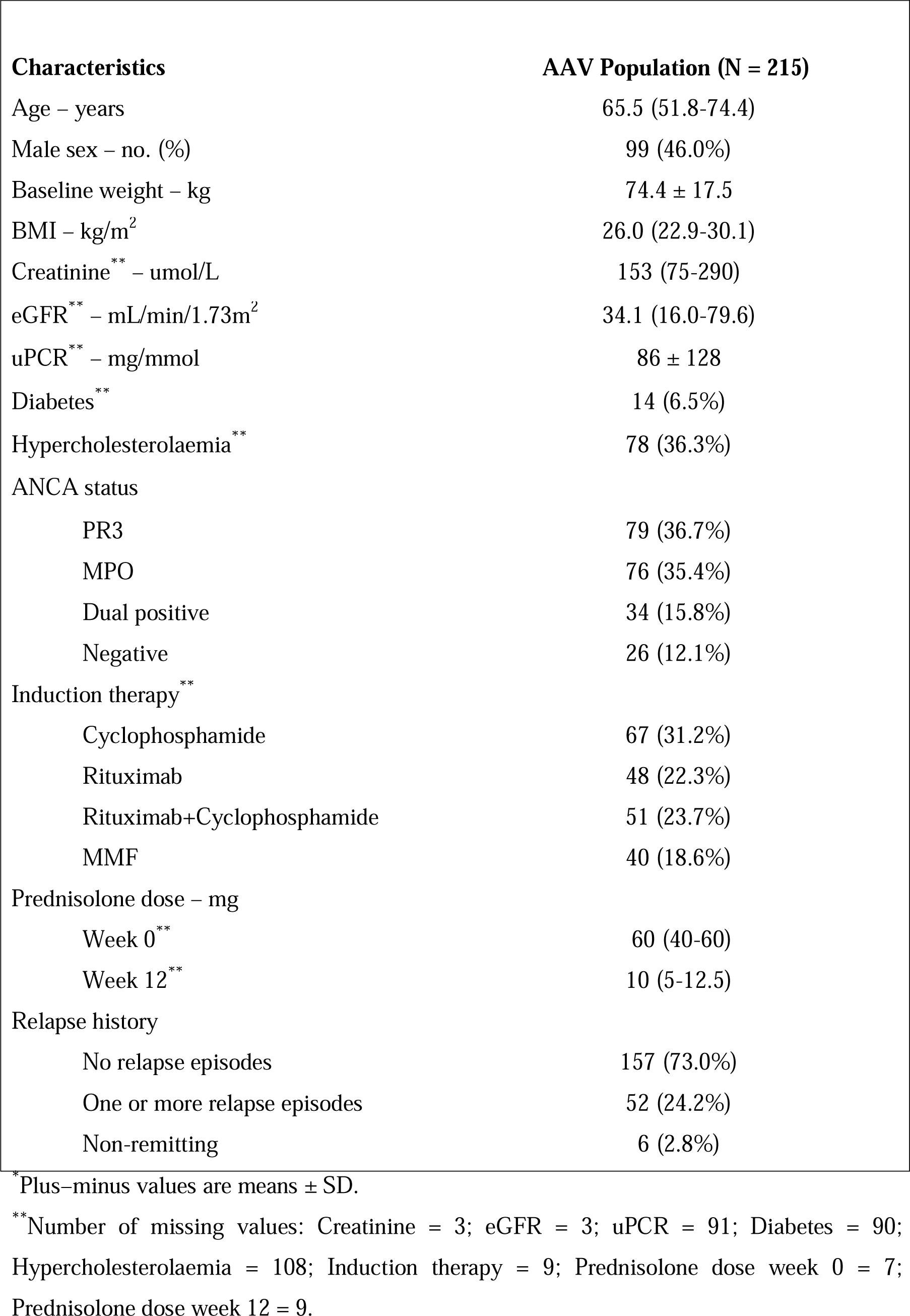
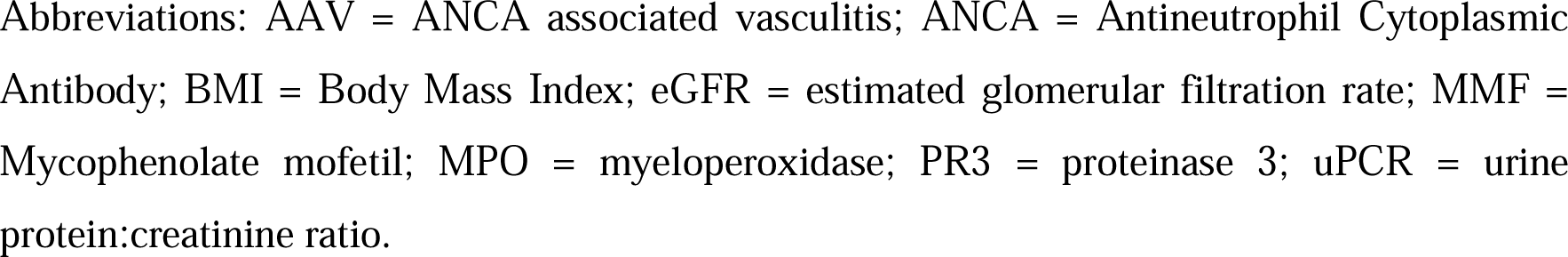
Demographic, Clinical & Laboratory Characteristics of Patients at Baseline.*

| Characteristics | AAV Population (N = 215) |
| --- | --- |
| Age – years | 65.5 (51.8-74.4) |
| Male sex – no. (%) | 99 (46.0%) |
| Baseline weight – kg | 74.4 ± 17.5 |
| BMI – kg/m<sup>2</sup> | 26.0 (22.9-30.1) |
| Creatinine** – umol/L | 153 (75-290) |
| eGFR** – mL/min/1.73m<sup>2</sup> | 34.1 (16.0-79.6) |
| uPCR** – mg/mmol | 86 ± 128 |
| Diabetes** | 14 (6.5%) |
| Hypercholesterolaemia** | 78 (36.3%) |
| ANCA status |  |
| PR3 | 79 (36.7%) |
| MPO | 76 (35.4%) |
| Dual positive | 34 (15.8%) |
| Negative | 26 (12.1%) |
| Induction therapy** |  |
| Cyclophosphamide | 67 (31.2%) |
| Rituximab | 48 (22.3%) |
| Rituximab+Cyclophosphamide | 51 (23.7%) |
| MMF | 40 (18.6%) |
| Prednisolone dose – mg |  |
| Week 0** | 60 (40-60) |
| Week 12** | 10 (5-12.5) |
| Relapse history |  |
| No relapse episodes | 157 (73.0%) |
| One or more relapse episodes | 52 (24.2%) |
| Non-remitting | 6 (2.8%) |

\*Plus-minus values are means ± SD.
\*\*Number of missing values: Creatinine = 3; eGFR = 3; uPCR = 91; Diabetes = 90; Hypercholesterolaemia = 108; Induction therapy = 9; Prednisolone dose week 0 = 7; Prednisolone dose week 12 = 9.
Abbreviations: AAV = ANCA associated vasculitis; ANCA = Antineutrophil Cytoplasmic Antibody; BMI = Body Mass Index; eGFR = estimated glomerular filtration rate; MMF = Mycophenolate mofetil; MPO = myeloperoxidase; PR3 = proteinase 3; uPCR = urine protein:creatinine ratio.

Baseline characteristics comparing PR3- and MPO-positive cohorts are shown in **Table 2**. PR3-AAV patients had a significantly higher baseline weight and BMI compared to MPO-AAV patients. PR3-AAV patients had better renal function at the time of diagnosis and were more likely to have received cyclophosphamide induction therapy.

**Table 2.** Comparison of Baseline Characteristics Between PR3 and MPO AAV.

| Characteristics | PR3 ANCA<br>(N = 79) | MPO ANCA<br>(N = 76) | P-value |
| --- | --- | --- | --- |
| Age – years | 57.4 ± 16.8 | 68.2 ± 14.4 | 0.18 |
| Male sex – no. (%) | 38 (48.1%) | 38 (50.0%) | 0.81 |
| Baseline weight – kg | 78.0 ± 20.9 | 70.4 ± 15.6 | 0.01 |
| BMI – kg/m<sup>2</sup> | 27.3 ± 6.3 | 24.9 ± 5.0 | 0.045 |
| Creatinine<sup>**</sup> – umol/L | 127 (71-234) | 196 (111.5-322.8) | 0.02 |
| eGFR<sup>**</sup> – mL/min/1.73m<sup>2</sup> | 46.2 (21.2-95.5) | 26.1 (13.9-50.9) | 0.003 |
| uPCR<sup>**</sup> – mg/mmol | 72 ± 102 | 94 ± 137 | 0.20 |
| Diabetes<sup>**</sup> | 2 (2.5%) | 6 (7.9%) | 0.13 |
| Hypercholesterolaemia<sup>**</sup> | 27 (34.2%) | 28 (36.8%) | 0.73 |
| Induction therapy |  |  |  |
| Cyclophosphamide | 33 (41.8%) | 19 (25.0%) | 0.03 |
| Rituximab | 16 (20.3%) | 22 (28.9%) | 0.21 |
| Rituximab+Cyclophosphamide | 18 (22.8%) | 23 (30.3%) | 0.29 |
| MMF | 10 (12.7%) | 11 (14.5%) | 0.74 |
| Prednisolone dose – mg |  |  |  |
| Week 0<sup>**</sup> | 60 (40-60) | 60 (40-60) | 0.76 |
| Week 12<sup>**</sup> | 10 (5-12.5) | 10 (5-12.5) | 0.09 |

\*Plus-minus values are means ± SD.
\*\* Number of missing values:
PR3 ANCA subgroup: Diabetes = 38; Hypercholesterolaemia = 48; Prednisolone dose week 0 = 4; Prednisolone dose week 12 = 4.
MPO ANCA subgroup: Creatinine = 2; eGFR = 2; Diabetes = 29; Hypercholesterolaemia = 34.
Abbreviations: AAV = ANCA associated vasculitis; ANCA = Antineutrophil Cytoplasmic Antibody; BMI = Body Mass Index; eGFR = estimated glomerular filtration rate; MMF = Mycophenolate mofetil; MPO = myeloperoxidase; PR3 = proteinase 3; uPCR = urine protein:creatinine ratio.

### Body weight trajectory after AAV diagnosis or relapse

Most patients (62.8%) gained weight during the first six months following AAV diagnosis and sustained this weight gain for at least two years (**Figure 1A**). On average, patients gained a mean 5.2 ±8.2% (P=0.001) of body weight by six months. During this period, a BMI increase of ≥5% was experienced by 87 patients (47.3%) and the median (±IQR) BMI increased to 27.5 (23.6-31.4) kg/m^2^ (+1.5 [0.7-1.3] kg/m^2^) with 61 patients (33.1%) documented as being overweight and 57 (31.0%) obese. Given this trajectory, we selected percentage change in BMI at 6 months as an appropriate end-point in the linear regression analyses presented below; particularly due to the acknowledged and clinically meaningful effects of weight fluctuations falling within the 3-5% range on both overall wellbeing and cardiovascular risk [13, 14]. The changes in individuals’ BMI categories from baseline to six months are outlined in supplemental Figure S1.

**Figure 1A:**
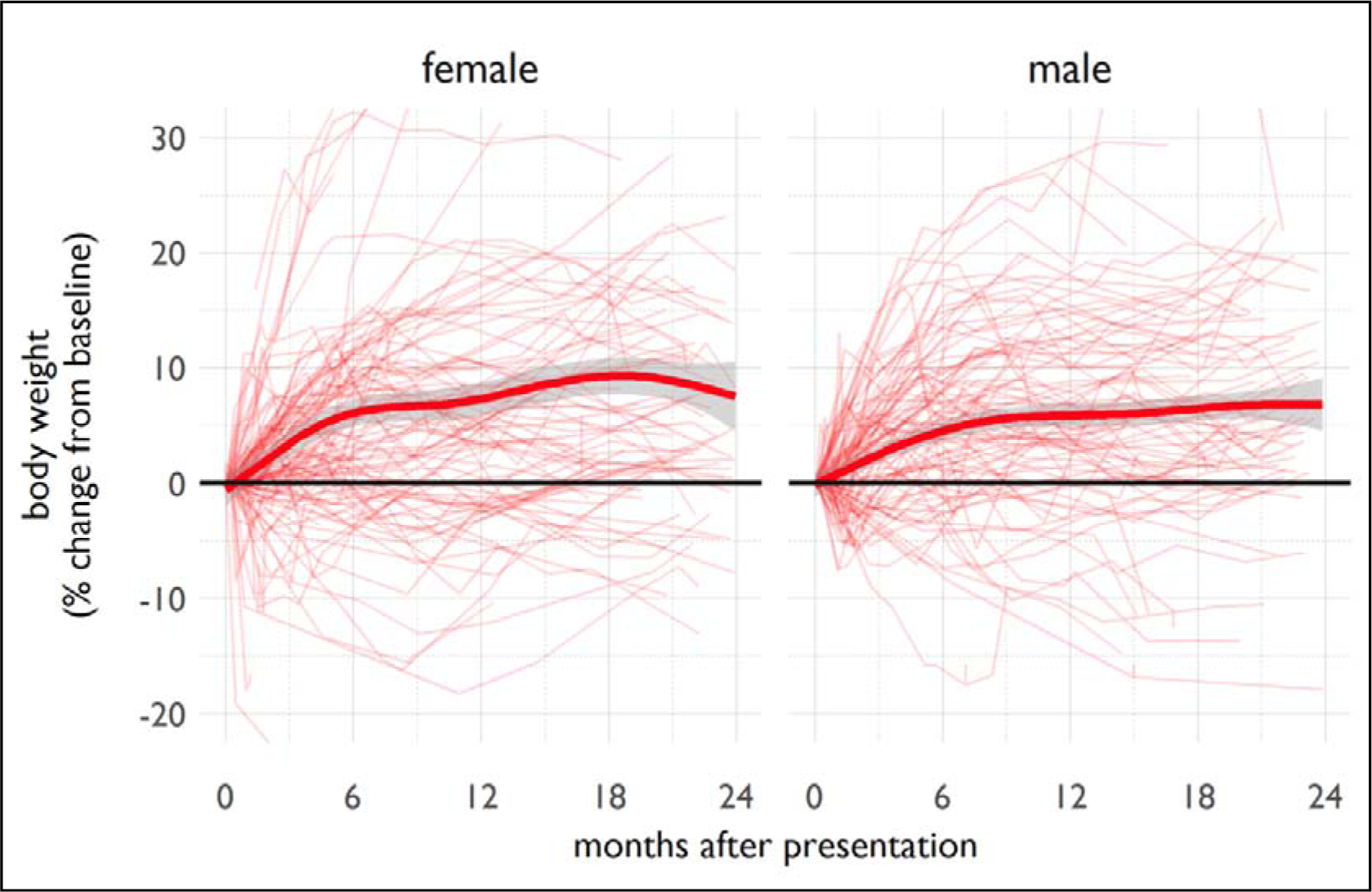
Trends in body weight following first presentation with AAV. Body weight for individuals plotted relative to body weight at time of presentation with vasculitis or first episode of disease relapse. The heavy line is the population average (smoothed conditional mean computed using a generalised additive model).

Weight gain following the first episode of disease relapse showed a less pronounced trend and reached its peak later after treatment (**Figure 1B**). Lower glucocorticoid doses were administered during disease relapse compared with index presentation treatment with median ±IQR prednisolone doses of 40 (20-40) mg and 7.5 (5-10) mg given at weeks zero and 12, respectively.

**Figure 1B:**
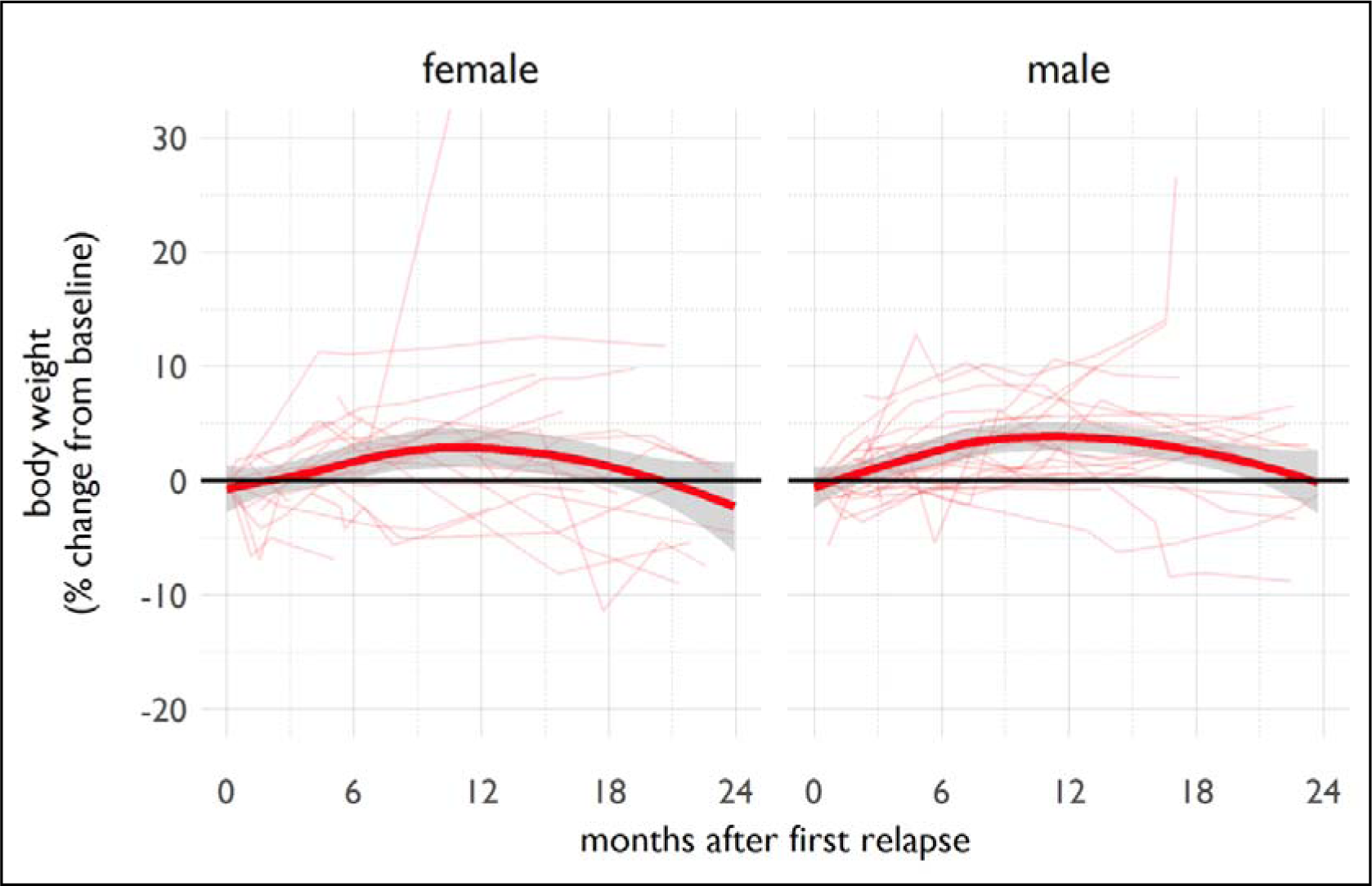
Trends in body weight following first episode of relapse of AAV. Body weight for individuals plotted relative to body weight at time of presentation with vasculitis or first episode of disease relapse. The heavy line is the population average (smoothed conditional mean computed using a generalised additive model).

### Risk factors for weight gain

Baseline characteristics associated with change in BMI at six months were a higher baseline eGFR (β=0.70 [0.36-1.03], P<0.001) and an earlier year of presentation (β=0.38 [0.08-0.69], P=0.008) (**Table 3**). A 10 mL/min/1.73m^2^ increase in eGFR corresponded to a BMI increase of 0.70% at six months from baseline. Those with an eGFR of >60 mL/min/1.73m^2^ experienced the greatest rise in BMI, with significant inter-individual variability (**Figure 2A)**. Additionally, for each year that the presentation date was in advance of the study’s conclusion there was a corresponding 0.38% increase in BMI (**Figures 2B**). The adjusted R^2^ value for the adjusted model was 0.102, meaning that 10.2% of the observed variation in BMI percentage change was explained by the linear regression model.

**Figure 2A:**
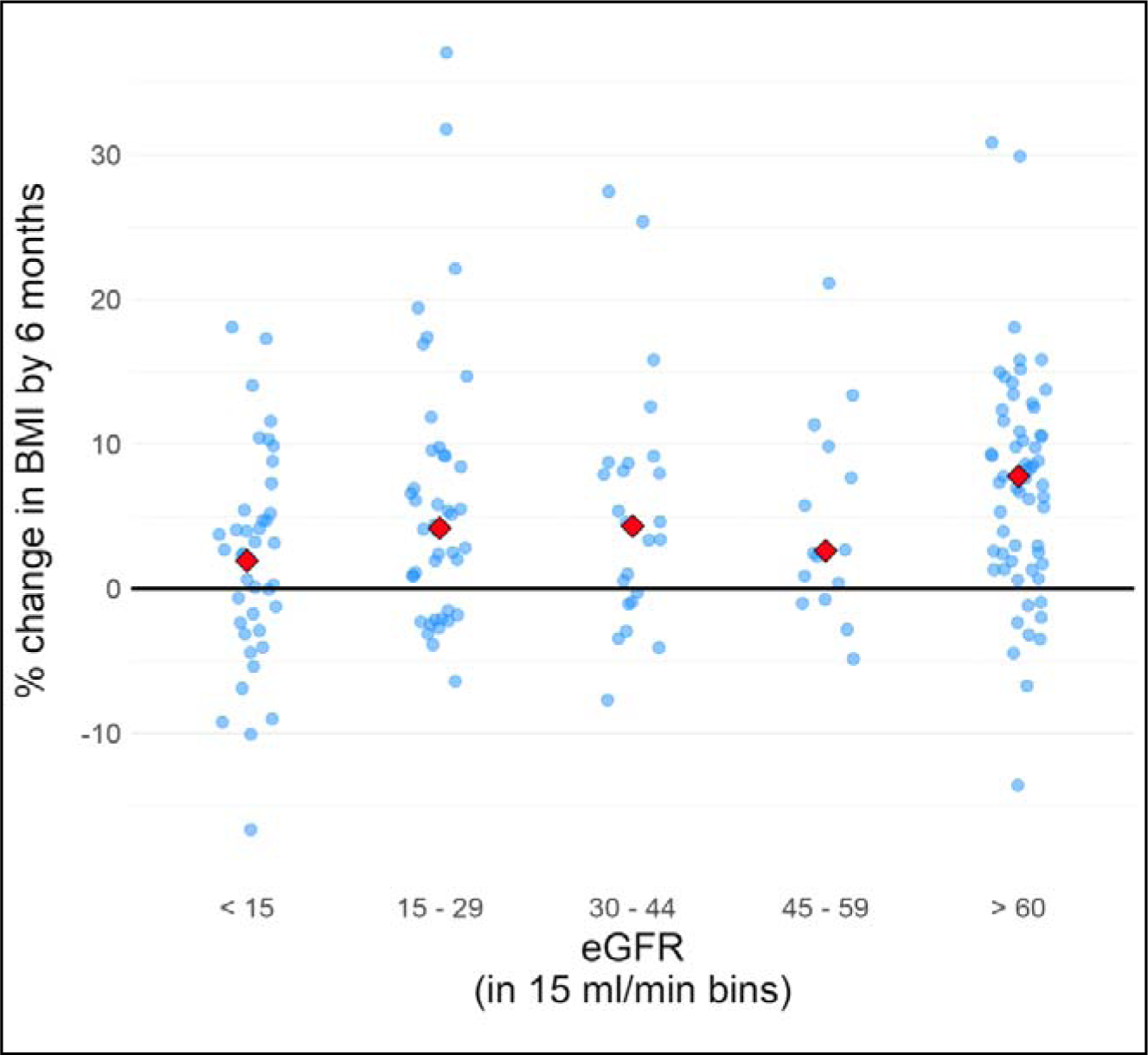
Percentage change in BMI experienced by patients at six months, grouped by eGFR.

**Figure 2B:**
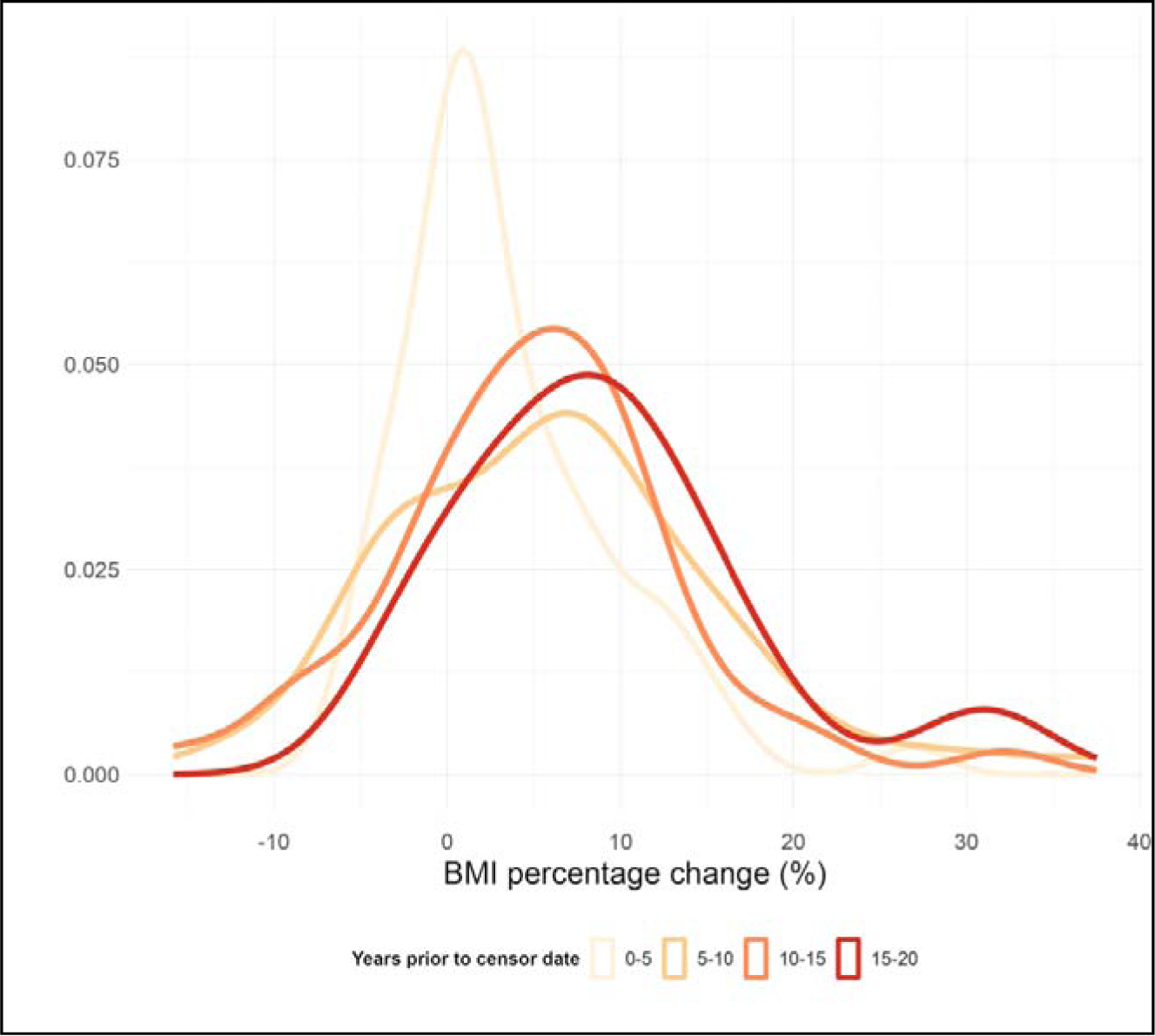
Percentage change in BMI experienced at six months, grouped by year of presentation to the vasculitis serv

**Table 3.** Determinants of BMI Percentage Change at Six Months.

| Characteristic | Unadjusted regression coefficients |  |  | Adjusted* regression coefficients |  |  |
| --- | --- | --- | --- | --- | --- | --- |
|  | Beta | 95% CI | P-value | Beta | 95% CI | P-value |
| Year of presentation | 0.22 | 0.01-0.44 | 0.039 | 0.38 | 0.08-0.69 | 0.008 |
| Age (years) | 0.34 | 0.19-1.74 | 0.084 | 0.29 | -0.15-0.73 | 0.35 |
| Age<sup>2</sup> (years<sup>2</sup>) | -0.05 | -0.01-0.00 | 0.077 | -0.01 | -0.01-0.00 | 0.52 |
| Sex |  |  |  |  |  |  |
| Male | 0 (ref) |  |  | 0 (ref) |  |  |
| Female | 1.15 | -0.83-3.13 | 0.253 | 0.77 | -1.27-2.77 | 0.45 |
| BMI at baseline | -0.10 | -0.26-0.07 | 0.245 | -0.17 | -0.35-0.01 | 0.06 |
| eGFR (10 mL/min/1.73m<sup>2</sup>) | 0.37 | 0.09-0.64 | 0.009 | 0.70 | 0.36-1.03 | <0.001 |
| uPCR | -0.002 | -0.01-0.00 | 0.380 | 0.00 | -0.01 -0.01 | 0.58 |
| Diabetes | -1.28 | -5.08-2.53 | 0.508 | -0.72 | -5.05-3.62 | 0.74 |
| Hyperlipidaemia | -1.40 | -4.58-1.79 | 0.385 | -2.53 | -5.83-0.78 | 0.13 |
| ANCA subgroup |  |  |  |  |  |  |
| Negative | 0 (ref) |  |  | 0 (ref) |  |  |
| MPO | -0.10 | -2.16-1.96 | 0.925 | 0.52 | -1.69-2.73 | 0.64 |
| PR3 | 1.22 | -0.81-3.25 | 0.238 | 0.75 | -1.52-3.01 | 0.52 |
| Dual positive | -0.75 | -3.51-2.01 | 0.594 | -0.91 | -3.80-1.97 | 0.53 |
| Induction agent |  |  |  |  |  |  |
| Cyclophosphamide | 1.68 | -0.42-3.78 | 0.116 | 0.83 | -2.51-4.17 | 0.63 |
| Rituximab | 0 (ref) |  |  | 0 (ref) |  |  |
| Rituximab+Cyclophosphamide | -0.68 | -3.02-1.66 | 0.565 | 0.04 | -0.71-5.58 | 0.13 |
| MMF | 0.07 | -2.37-2.53 | 0.951 | 2.44 | -3.22-3.30 | 0.98 |
| Prednisolone dose |  |  |  |  |  |  |
| Week 0 | 0.04 | -0.01-0.10 | 0.105 | 0.05 | -0.01-0.10 | 0.12 |
| Week 12 | 0.03 | -0.13-0.19 | 0.683 | -0.03 | -0.23-0.16 | 0.74 |

\* Adjusted for age, BMI at baseline, eGFR, induction regime and steroid dose at zero months.
Adjusted R<sup>2</sup> for final model: 10.2%
Abbreviations: ANCA = Antineutrophil Cytoplasmic Antibody; BMI = Body mass index; CI = Confidence interval; eGFR = estimated Glomerular filtration rate; MMF = Mycophenolate mofetil; MPO = myeloperoxidase; OR = Odds ratio; PR3 = proteinase 3; uPCR = Urine protein:creatinine ratio.

A higher baseline eGFR (aOR = 1.36 (1.08-2.72), p<0.001) and higher baseline BMI (aOR = 2.57 (1.81-3.64), P<0.001) conferred a significantly increased likelihood of obesity at six months from diagnosis (**Table 4**). Of the 56 (26.0%) individuals who were obese at baseline, 54 (25.1%) were also obese at six months. The area under the receiver-operator characteristic (ROC) curve for the adjusted regression model decreased from 0.97 to 0.70 when baseline BMI was not included in the logistic regression model, demonstrating that baseline BMI accounts for much of the predictive power of this model.

**Table 4.** Odds Ratios for Obesity at Six Months from Baseline.

| Characteristic | OR | 95% CI | P-value | aOR<sup>1</sup> | 95% CI | P-value |
| --- | --- | --- | --- | --- | --- | --- |
| Year of presentation | 1.02 | 0.96-1.09 | 0.52 | 1.09 | 0.88-1.34 | 0.43 |
| Age (deciles) | 0.79 | 0.65-0.97 | 0.02 | 1.66 | 0.94-2.93 | 0.08 |
| Sex |  |  |  |  |  |  |
| Male | 1.00 (ref) | NA | NA | 1.00 (ref) | NA | NA |
| Female | 0.84 | 0.45-1.55 | 0.57 | 0.92 | 0.23-3.70 | 0.91 |
| BMI at baseline | 2.08 | 1.66-2.60 | <0.001 | 2.57 | 1.81-3.64 | <0.001 |
| eGFR (10 mL/min/1.73m<sup>2</sup>) | 1.09 | 1.00-1.19 | 0.04 | 1.36 | 1.08-2.72 | <0.001 |
| uPCR | 1 | 0.99-1.00 | 0.95 | 1.00 | 0.99-1.00 | 0.51 |
| Diabetes | 1.32 | 0.37-4.77 | 0.67 | 1.11 | 0.25-4.99 | 0.89 |
| Hyperlipidaemia | 0.48 | 0.18-1.28 | 0.14 | 0.45 | 0.11-1.86 | 0.27 |
| ANCA subgroup |  |  |  |  |  |  |
| Negative | 1.00 (ref) | NA | NA | 1.00 (ref) | NA | NA |
| MPO | 0.46 | (0.23-0.90) | 0.02 | 0.33 | 0.02-4.92 | 0.42 |
| PR3 | 2.09 | 1.12-3.93 | 0.02 | 1.11 | 0.11-10.9 | 0.93 |
| Dual positive | 1.05 | 0.46-2.43 | 0.9 | 0.26 | 0.02-3.40 | 0.30 |
| Induction agent |  |  |  |  |  |  |
| Cyclophosphamide | 0.84 | 0.43-1.62 | 0.6 | 0.4 | 0.06-2.89 | 0.36 |
| Rituximab | 1.00 (ref) | NA | NA | 1.00 (ref) | NA | NA |
| Rituximab+Cyclophosphamide | 0.58 | 0.30-1.25 | 0.16 | 0.84 | 0.09-8.08 | 0.88 |
| MMF | 1.68 | 0.80-3.51 | 0.17 | 0.25 | 0.03-2.41 | 0.23 |
| Prednisolone dose |  |  |  |  |  |  |
| Week 0 | 1.02 | 1.00-1.04 | 0.02 | 0.99 | 0.94-1.04 | 0.67 |
| Week 12 | 1.03 | 0.98-1.08 | 0.27 | 1.01 | 0.88-1.17 | 0.89 |

\* Adjusted for age, gender, baseline BMI, eGFR, induction agent, prednisolone dose at baseline and 12 weeks, and ANCA subgroup
Abbreviations: aOR = Adjusted odds ratio; ANCA = Antineutrophil Cytoplasmic Antibody; BMI = Body mass index; CI = Confidence interval; eGFR = estimated Glomerular filtration rate; MMF = Mycophenolate mofetil; MPO = myeloperoxidase; OR = Odds ratio; PR3 = proteinase 3; uPCR = Urine protein:creatinine ratio.

### Association between obesity and relapse rate

In order to test for an association between obesity and relapse rate, we performed a multivariate cox proportional hazard regression (supplemental Table S1). A baseline BMI of ≥30 kg/m^2^ was not associated with an increased risk of relapse after adjusting for confounding variables (HR P=0.85) (supplemental Figure S2). The cumulative relapse hazard curves of obese and non-obese patients did overlap (log-rank P<0.001).

## Discussion

### Individuals gain weight after a diagnosis of AAV

In a real-world setting, we have shown that individuals tend to experience large gains in body weight after a diagnosis of AAV. Similar to a clinical trial population [4], we found that weight gain typically occurs within the first six months after diagnosis. By six months, median BMI had increased by 1.5 units, nearly half of the patients had experienced a ≥5% rise in BMI and two-thirds of patients were either overweight or obese. Importantly, we found that changes in body weight and BMI were sustained for years. This is expected to have significant deleterious health consequences. A 5% gain in weight is associated with clinically meaningful increases in the risks of cardiovascular and metabolic disease [13, 14]. It is also associated with reductions in quality of life [15-17].

We found that the extent of weight gain observed following the first episode of disease relapse was less pronounced, echoing findings from the RAVE trial [4]. Our steroid-prescribing data suggest that this is likely to reflect reduced glucocorticoid exposure during treatment of relapse (*versus* first presentation). It is possible that this is also explicable by a shorter diagnostic delay, limiting the duration of any hypercatabolic state causing weight loss prior to treatment initiation.

### Risk factors for weight gain

In our cohort, the baseline factors associated with an increase in BMI included better eGFR and disease presentation in an earlier calendar year. A higher baseline eGFR was associated with an increased odds of being obese at six months. The apparently protective effects of lower baseline eGFR against BMI rise may be due to the anorexic effects of uraemia and/or more conservative glucocorticoid dosing in patients with advanced kidney disease in whom there may been consideration of treatment futility. The small R^2^ in our analyses supports the presence of other confounding factors, not adjusted by selected variables in our regression models.

We did not find a significant association between glucocorticoid exposure and elevated BMI. This should not be interpreted as evidence that glucocorticoids are not the predominant drivers of weight gain. Indeed, it is overwhelmingly likely that the weight gain experienced after AAV diagnosis is predominantly driven by glucocorticoid therapy, especially given the observed improvement in degree of weight gain in more contemporary treatment eras within our cohort. The lack of any association likely reflects a lack of variation in glucocorticoid dose at the observed time-points. (There is an analogy with some early epidemiological studies of lung cancer; to paraphrase Richard Peto, “if everybody smoked then lung cancer would be a genetically-determined disease”). Current treatment regimens use relatively low doses of glucocorticoids, which have proven to be effective and safe compared to historic standard-dose regimens [18, 19]. Rapid glucocorticoid withdrawal has proven to be effective and safe in smaller scale studies [20], and glucocorticoid–free rituximab monotherapy has good outcome data to now be recommended as first line maintenance therapy agent in AAV treatment [21-23]. Novel steroid-sparing agents such as oral C5a inhibitors, may be able to further reduce glucocorticoid-related toxicity [24]. Our observation that calendar year of presentation was associated with BMI changes, with greater weight gain for patients presenting longer ago, is likely explained by a trend towards more conservative steroid prescribing over the two decades spanned by our study). The changes in our centre’s immunosuppression therapy over time, specifically induction regimens and glucocorticoid dosages, are reflected in supplemental Figures S3 and S4.

Unsurprisingly, high BMI at presentation was a strong risk factor for high BMI at six months. Therefore, baseline BMI may be a useful parameter to include when prioritising individuals for steroid-sparing therapies (e.g., C5a inhibitors). ANCA serotype was not associated with a differential rate of weight gain. However, our observations align with previous published literature, indicating that PR3-AAV patients present with higher baseline body weight and BMI compared to MPO-AAV [10], potentially placing this AAV subgroup at greater risk of unhealthy high weight following treatment. In keeping with the literature [25, 26], MPO positive patients in our cohort exhibited inferior renal function at the time of diagnosis. The lower baseline BMI we observed within this subgroup could potentially be explained by the often-delayed diagnosis of MPO-AAV resulting in a more prolonged period of constitutional symptoms and uraemia.

### Association between obesity and relapse risk

We did not detect an association between obesity and risk of AAV disease relapse. This is noteworthy because it has been hypothesised that the relapsing nature of PR3-AAV might be exacerbated by the pro-inflammatory state induced by baseline obesity [10, 25].

### Strengths and limitations

We provide the most comprehensive characterisation to date of changes in body weight and BMI following a diagnosis of AAV, in a contemporary ‘real world’ cohort.

There are several limitations, in addition to the general limitations inherent in an observational study. Our population is predominantly White Caucasian, limiting the generalisability of our findings. We were not able to quantify the contribution to weight gain from glucocorticoids because of a lack of variation in glucocorticoid dosing. More than half of the cohort (N = 119) received 60 mg of prednisolone at treatment onset and only one third (N = 67) had their dose tapered to 10 mg at three months, potentially resulting in a model that underestimated the differential effect of glucocorticoids on weight gain. This question would be better addressed by future large-scale prospective studies in an era of lower – and more variable – glucocorticoid use or in RCTs.

### Conclusion and perspective

In the active phase of AAV, patients commonly experience a hypercatabolic state and weight loss, contributing to poorer outcomes [27, 28]. However, treatment often leads to weight gain, posing risks of future obesity and metabolic syndrome, which increases cardiovascular risk and reduces quality of life [1, 9, 29, 30]. Patients frequently report fatigue, pain, depression, anxiety, and work disability [31-33]. The aetiology of these symptoms remains poorly understood, yet fluctuations in body weight may play a role.

Our study corroborates that weight gain and obesity are common problems for patients with AAV. The substantial majority of patients in our cohort were overweight or obese by 6 months after diagnosis. We found that major risk factors for weight gain were higher baseline renal function and earlier eras of disease diagnosis (with presumed greater cumulative glucocorticoid exposure). These factors – combined with baseline BMI – could be used to identify patients at risk of becoming overweight, to prioritise individuals for steroid-sparing treatment regimens and pro-active weight management intervention. Such intervention would be multimodal, including lifestyle changes, conventional cardiovascular risk reduction strategies [34] and possibly novel pharmacological agents promoting weight loss and cardiovascular protection such as sodium-glucose cotransporter-2 (SGLT-2) inhibitors and GLP1 agonists [35, 36].

## Article information

### Data availability statement

The data that support the findings of this study are available from the corresponding author [TS] upon request and subject to our being able to satisfy data-sharing agreements that preserve patient confidentiality.

## Data Availability

All data produced in the present study are available upon reasonable request to the authors. The code used for analysis of the presented data is publicly available.

https://github.com/TWFrench/ANCA_vasculitis/tree/main

