## Supplemental Appendix for "Weight Gain Following a Diagnosis of Anti-neutrophil Cytoplasm Antibody-Associated Vasculitis"

**Supplementary Appendix**

The code used for analysis of the presented data is publicly available in the following repository: https://github.com/TWFrench/ANCA_vasculitis/tree/main.

**Figure S1** illustrates the changes in BMI categories among individuals from baseline to six months following diagnosis of AAV.

The results of the multivariate cox proportional hazard regression are displayed in **Table S1**. A baseline BMI of ≥30 kg/m^2^ was not associated with an increased risk of relapse after adjusting for confounding variables (HR 1.07 [0.53-2.16], P=0.846). Kaplan-Meier curves comparing risk of relapse in the obese cohort and the not-obese cohort did not overlap (log-rank P<0.001) (**Figure S2**). Increasing age (HR 1.57 [1.18-2.09], P=0.002), higher baseline eGFR (HR 1.19 [1.08-1.31], P<0.001) and use of cyclophosphamide rather than rituximab (HR 4.43 [1.53-12.8], P=0.006), conferred a greater risk of experiencing one or more relapse episodes in the follow-up period. Rates of relapse in the population treated with cyclophosphamide alone were significantly higher than those in the population treated with any one of the three alternative treatment regimens (log-rank P<0.001). The changes in our centre’s immunosuppression therapy over time, specifically induction regimens and glucocorticoid dosages, are reflected in **Figures S3** and **S4**. As depicted, cyclophosphamide monotherapy was predominantly utilised in earlier time periods in our study. However, we found that the risk of relapse following cyclophosphamide induction was irrespective of the year of presentation and less likely to be confounded by evolutions in glucocorticoid dosing regimens over time. **Figure S4** highlights prednisolone doses at presentation and three months and suggests a more rapid glucocorticoid tapering in recent years.

| Table S1. Hazard Ratios for Likelihood of Experiencing ≥1 Relapse Episodes. | | | |
| --- | --- | --- | --- |
| Characteristic | **Adjusted HR** | **95% CI** | **P-value** |
| Baseline obesity | 1.07 | 0.53 -2.17 | 0.846 |
| Presentation year | 0.99 | 1.18-2.09 | 0.867 |
| Age (deciles) | 1.57 | 1.08-1.31 | 0.002 |
| Female sex | 0.98 | 0.55-1.75 | 0.94 |
| eGFR (10 mL/min) | 1.19 | 0.90-1.09 | <0.001 |
| ANCA sub-group^*^ |  |  |  |
| PR3 | 3.08 | 0.89-10.6 | 0.075 |
| MPO | 0.62 | 0.15-2.51 | 0.503 |
| Dual positive | 1.07 | 0.26-4.43 | 0.923 |
| Induction agent^**^ |  |  |  |
| Cyclophosphamide | 4.43 | 1.53-12.8 | 0.006 |
| Rituximab +Cyclophosphamide | 0.28 | 0.03-2.42 | 0.247 |
| MMF | 2.43 | 0.77-7.80 | 0.136 |

^*^Relative to ANCA negative
^**^Relative to induction with rituximab

Abbreviations: ANCA = Anti-neutrophil Cytoplasm Antibody; CI = Confidence interval; eGFR = Estimated glomerular filtration rate; HR = Hazard ratio; MMF = Mycophenolate mofetil; MPO = Myeloperoxidase; PR3 = Proteinase 3; uPCR = Urine protein:creatinine ratio.

**Figure S1.** Changes in BMI category from baseline to six months following diagnosis of AAV.

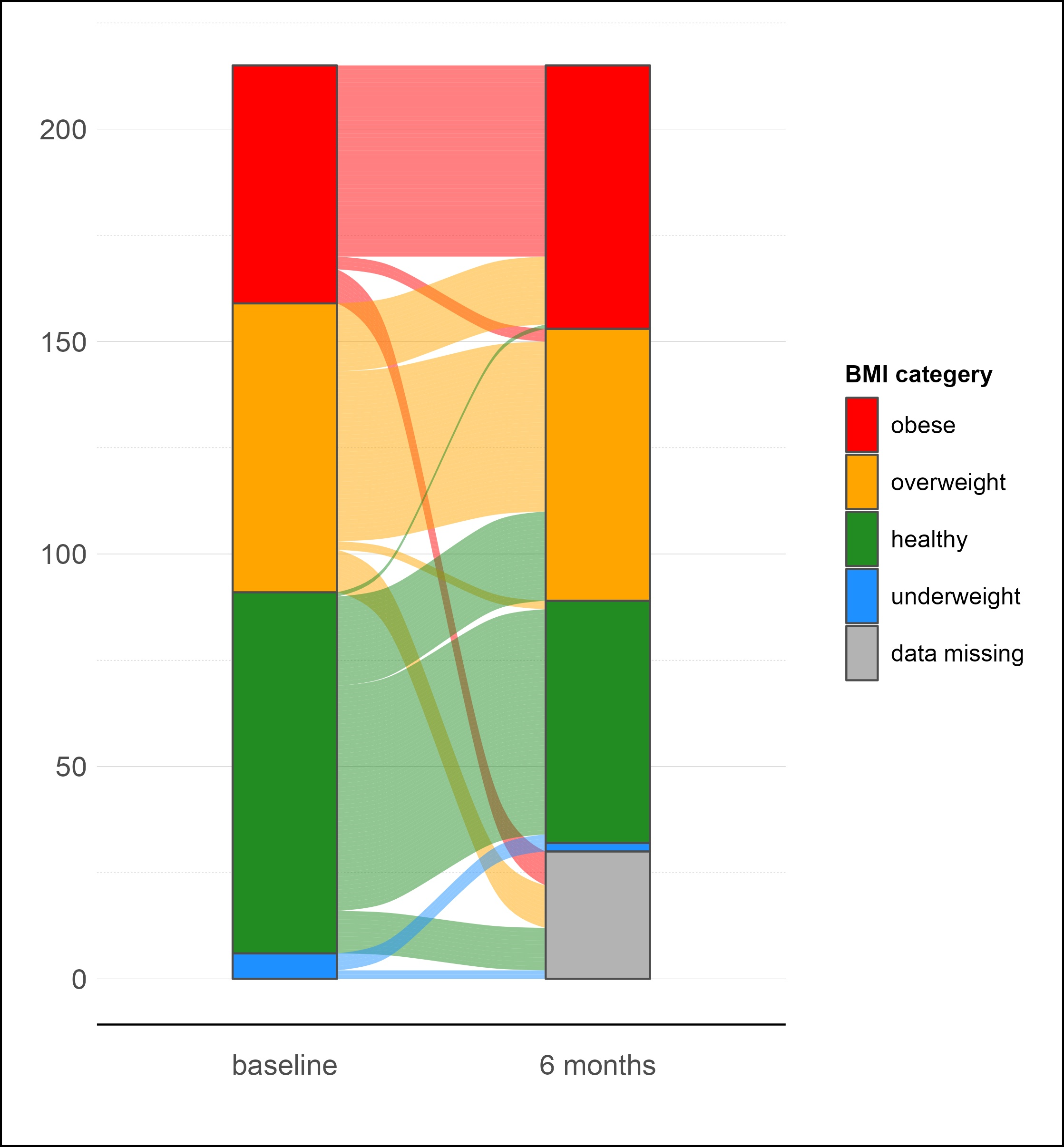

**Figure S2.** Kaplan-Meier plot on cumulative risk of relapse over time following initial presentation in obese and non-obese participants. Obese group in black, comparison group in red. Log-rank test P-value <0.900.

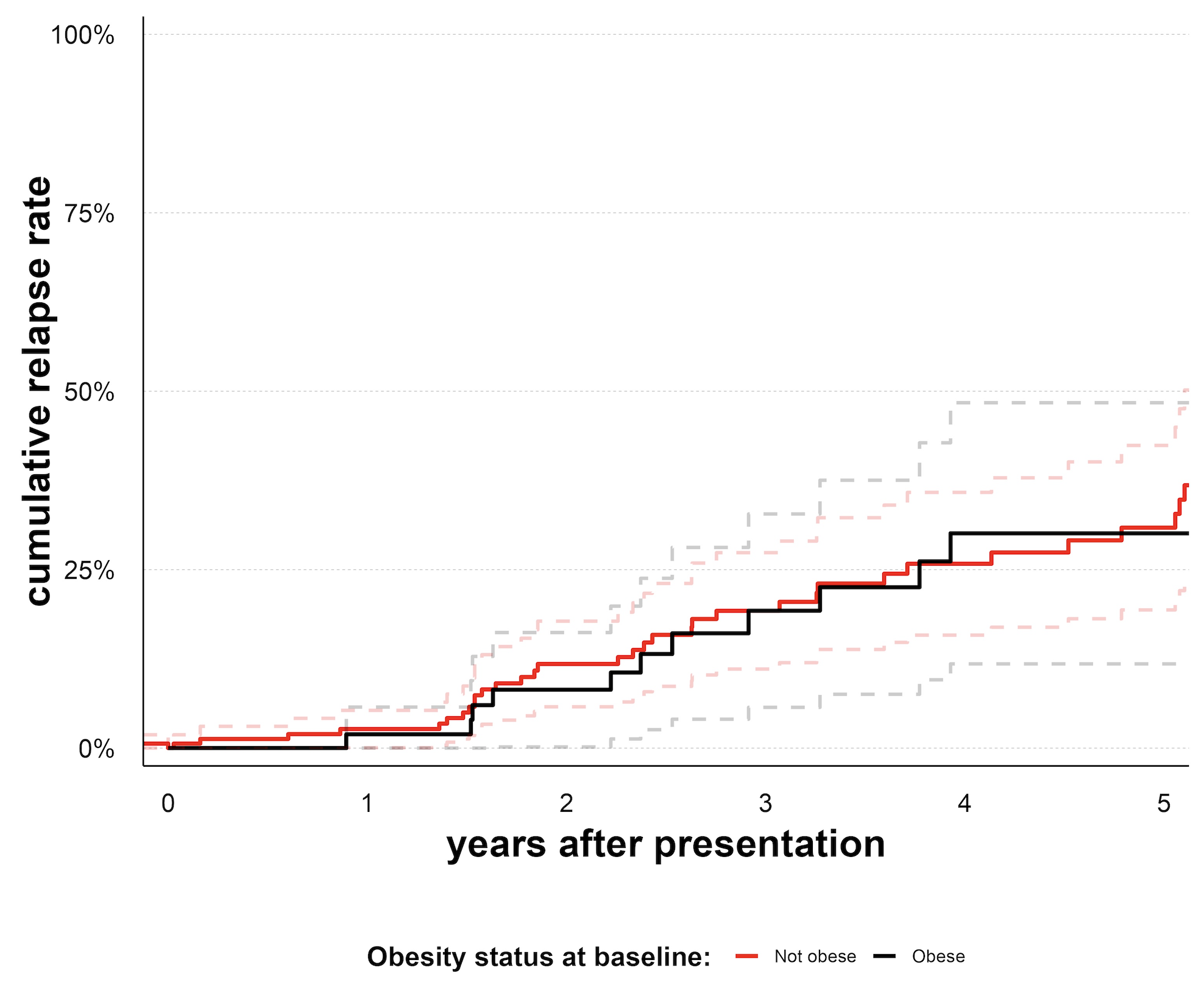

**Figure S3.** Percentage plot demonstrating changes in induction immunosuppression agents over time.

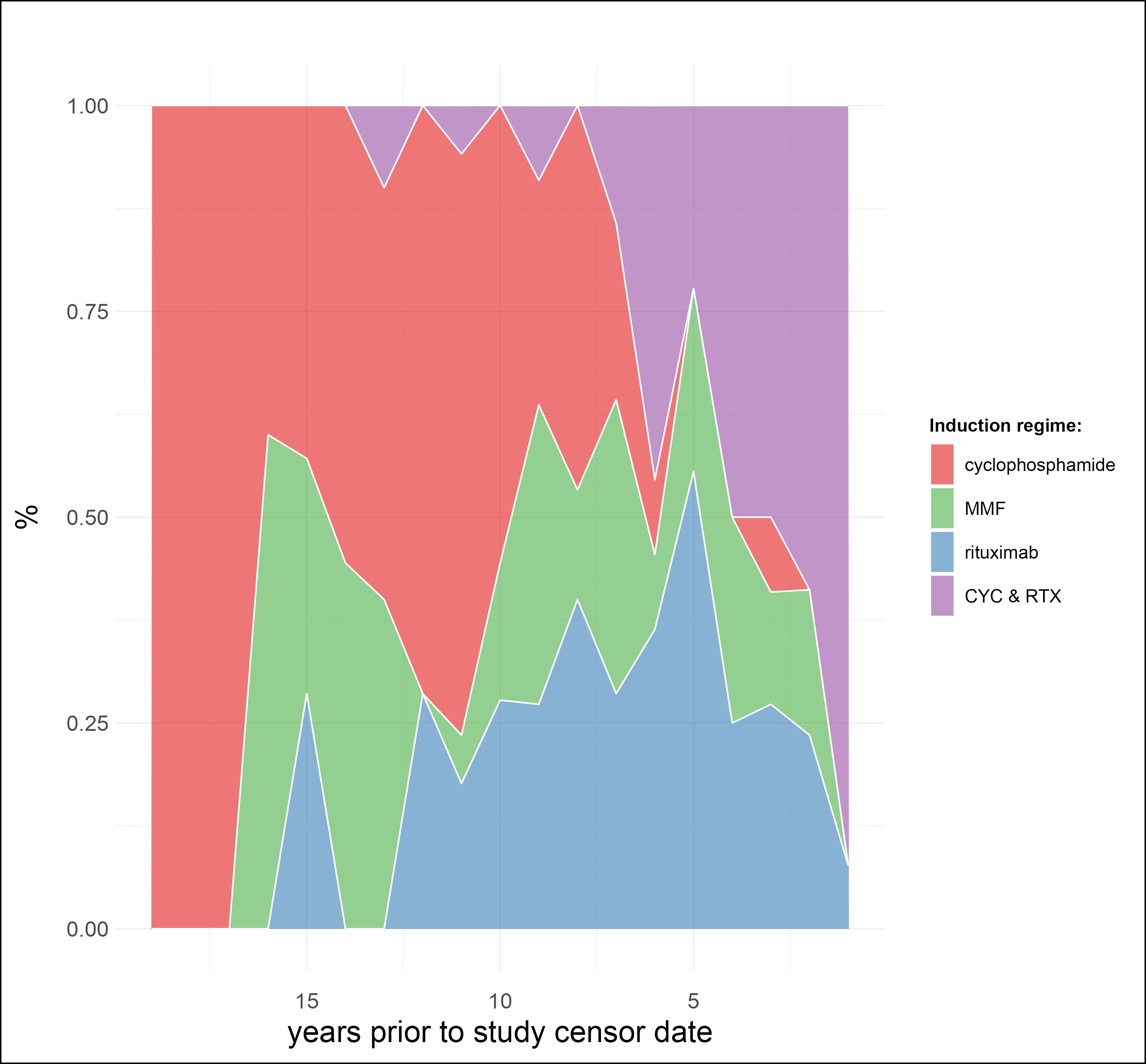

**Figure S4.** Prednisolone doses at presentation and three months, grouped by year of presentation to the vasculitis service.

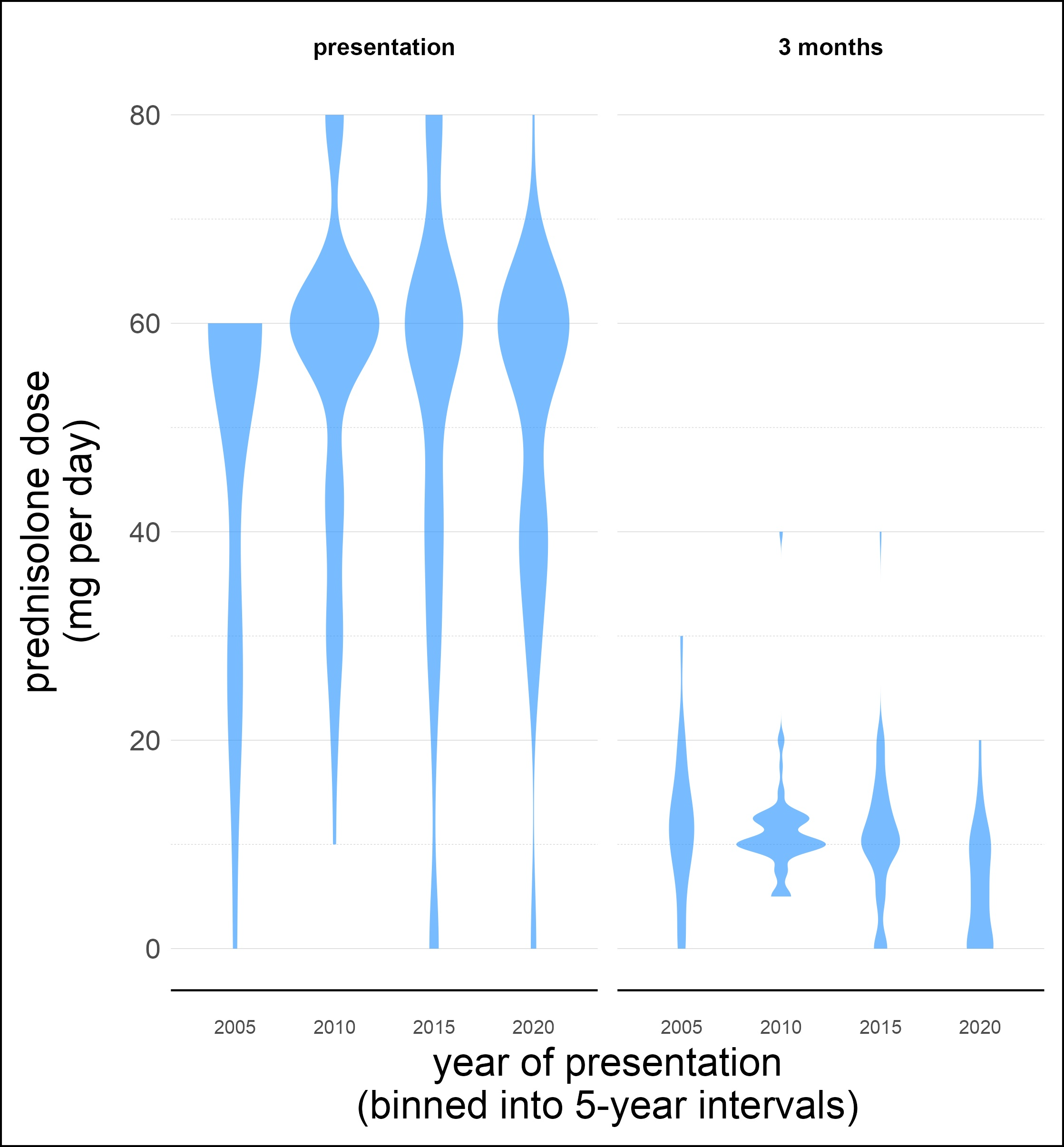
